# Machine Learning-Based Prediction of Pediatric Ulcerative Colitis Treatment Response using Diagnostic Histopathology

**DOI:** 10.1101/2024.01.22.24301559

**Authors:** Xiaoxuan Liu, Thomas Walters, Iram Siddiqui, Oscar Lopez-Nunez, Surya Prasath, Lee A Denson, PROTECT consortium, Jasbir Dhaliwal

## Abstract

**Background and Aims:** We previously reported clinical features associated with outcomes in pediatric ulcerative colitis (UC). Here we developed a histopathology model to predict corticosteroid-free remission (CSFR) on mesalamine therapy alone.

**Methods:** Pre-treatment rectal biopsy slides were digitized in training and validation groups of 292 and 113 pediatric UC patients, respectively. Whole slide images (WSI) underwent pre-processing. Thirteen machine learning (ML) models were trained using 250 histomic features including texture, color, histogram, and nuclei. Feature importance was determined by the Gini index with the classifier re-trained using the top features.

**Results:** 187571 informative patches from 292 training group patients (Male:53%; Age:13y (IQR:11-15); CSFR:41%) were trained on 13 ML classifiers. The best model was random forest (RF). Eighteen optimal histomic features were identified and trained, and the corresponding WSI AUROC was 0.89 (95%CI:0.71, 0.96), accuracy of 90% for CSFR. Features were re-trained on an independent real-world dataset of 113 patients and the model WSI AUROC was 0.85 (95%CI:0.75, 0.95), accuracy of 85%.

**Conclusion:** Routine histopathology obtained at diagnosis contains histomic features associated with UC treatment response.

## INTRODUCTION

The initial presentation of ulcerative colitis (UC) within the pediatric population exhibits a degree of uniformity, the majority characterized by extensive colitis at time of diagnosis. However, the response to initial therapy demonstrates marked heterogeneity.^1^ It is challenging to discriminate which patients would successfully improve on corticosteroids followed by mesalamine (5-ASA therapy) maintenance therapy, and those who would benefit from early introduction of biologic therapy. Identifying the optimal temporal window for the administration of biologic treatments and effective stratification of patients based on associated risk factors, remains a pivotal yet an unmet clinical exigency.

The clinical and biological predictors of response to standardized pediatric colitis therapy (PROTECT) study, a multicenter study of newly diagnosed UC children, aimed to address this, primarily identifying those children that would be successfully maintained on mesalamine (5-ASA) maintenance therapy at one year.^1^ The study identified three early clinical features, pediatric ulcerative colitis activity index (PUCAI) <45, hemoglobin ≥10g/dL at time of diagnosis and week 4 clinical remission as predictors of steroid-free clinical remission. The resultant predictive model demonstrated an area under the curve of 0.70 (95%CI 0.65, 0.75) and cross validation of 0.63 (95%CI 0.57, 0.69). Moreover, PROTECT offered novel insights into the prognostic utility of histological features assessed at disease onset, notably surface villiform architectural abnormality^1,2^ and rectal eosinophilia. The predictive capacity of histological features has been most recently evaluated in the setting of clinical relapse and treatment response^3, 4^, but there is a paucity of studies evaluating the prognostic role of such features at diagnosis.

The manual evaluation of histological slides remains indispensable. However, in the setting of a predictive tool that has the potential for wide adoption, such an approach would be restrictive. Manual evaluation is time consuming and the reported inter- and intra-rater variability ranging from 0.48 to 0.94^5^. Alternatively, automated image processing can provide standardized quantitative and high-throughput analysis that has the potential to be widely implemented in clinical practice. In oncopathology image analysis frameworks have been developed to evaluate whole slide digital pathology images for classification tasks^6–8^, by extracting histomorphometric features from the images. These features otherwise known as histomic or handcrafted features, have been mathematically engineered to capture the underlying properties of the tissue^9–11^.

In this study we used the PROTECT diagnostic treatment naïve hematoxylin and eosin (H+E) stained rectal biopsies for whole slide imaging (WSI) and advanced computational approaches to develop an automated histomics based model to predict corticosteroid free clinical remission (CSFR) with the only therapy being mesalamine at one year following diagnosis.

## MATERIAL AND METHODS

### Study Participants

PROTECT was a multicenter inception cohort study based at 29 centers in the United States and Canada^12^. 400 children aged 4–17 years with a diagnosis of UC based on established clinical, endoscopic, and histological parameters were included. Inclusion criteria included disease extension beyond the rectum, a baseline Pediatric Ulcerative Colitis Activity Index (PUCAI) score of at least 10, no previous therapy for colitis, and stool culture negative for enteric bacterial pathogens, including Clostridium difficile toxin. Detailed protocol and study description can be found in Hyams et al^1, 12^. Depending on the initial PUCAI score (PUCAI less than 10 denoted inactive disease or remission, 10−30 denoted mild disease, 35−60 denoted moderate disease, and 65 or higher denoted severe disease), patients received initial treatment with either mesalamine (mild disease), or corticosteroids (moderate and severe disease), with physician discretion permitted. A detailed description of treatment guidelines is provided in Hyams et al^1^. All patients on mesalamine received study drug in the form of Pentasa (Shire Pharmaceuticals/Pantheon, Greenville, NC, USA).

In the current study, a sub-cohort of 292 treatment naïve patients with available rectal biopsy samples from the PROTECT study were included (Table 1) for model development. The external validation test cohort included 113 pediatric patients enrolled and prospectively followed in the Canadian Children IBD Network (CIDsCANN) inception cohort study at the Hospital for Sick Children (Sickkids), Toronto, one of 12 participating sites, (Supplementary Table 1).^13, 14^ The biopsy samples from both cohorts were taken from the most inflamed part of the rectosigmoid and were routinely processed and fixed in formalin and embedded in paraffin blocks from which 4–5 micron sections were cut and stained with hematoxylin and eosin (H+E) (Roche, HE600). All slides were scanned at 20x, Aperio T2 for digital analysis. The PROTECT biopsies were all processed at Cincinnati Children’s Medical Center, and Sickkids biopsies in their clinical pathology laboratory.

**Table 1:** Demographic clinical and histological characteristics of training cohort stratified by primary outcome of corticosteroid free remission on mesalamine alone at one year.

|  | Steroid free remission<br>N=118 | Not in steroid free<br>remission<br>N=174 | P-value |
| --- | --- | --- | --- |
| Age (years) | 13 (10, 15) | 13.5 (11, 15) | 0.59 |
| Race |  |  |  |
| White | 101 | 141 | 0.37 |
| Black | 6 | 11 |  |
| Asian | 7 | 5 |  |
| Male | 63 (53%) | 91 (52%) | 0.89 |
| Disease Extent†<br>Distal (E1/E2):<br>Extensive(E3/E4) | 31 (26%) : 87 (74%) | 26 (15%) : 148 (85%) | 0.05 |
| ***Disease activity at presentation*** |  |  |  |
| PUCAI†† | 45 (25, 60) | 55 (40, 70) | 0.0003 |
| Endoscopic Mayo |  |  |  |
| Mayo 1 | 22 (19%) | 16 (9%) | 0.002 |
| Mayo 2 | 70 (59%) | 88 (51%) |  |
| Mayo 3 | 26 (22%) | 70 (40%) |  |
| Nancy Index* |  |  |  |
| Grade 1 | 18(56%) | 14(44%) | 0.009 |
| Grade 2 | 74(44%) | 95(56%) |  |
| Grade 3 | 26(29%) | 65(71%) |  |
| ***Initial Induction therapy*** |  |  |  |
| Mesalamine | 56 (47%) | 42 (24%) | 0.0003 |
| Oral corticosteroids | 36 (31%) | 65 (37%) |  |
| IV corticosteroids | 26 (22%) | 67 (39%) |  |
| Expressed as median (interquartile range); †Disease extent: E1 ulcerative proctitis; E2 Left sided UC; E3 Extensive (hepatic flexure distally); E4 Pancolitis; ††PUCAI: Pediatric Ulcerative Colitis Activity Index; *Nancy index: Grade 1 none/chronic inflammation only; Grade 2 inclusive of acute inflammation without crypt abscesses; Grade 3 inclusive of abscess |  |  |  |

We used the PROTECT study primary outcome of corticosteroid-free remission (CSFR) at one year on mesalamine therapy alone and with no colectomy. Clinical remission was defined as a PUCAI score <10 and with no corticosteroid use for 4 weeks or longer immediately prior to one year. The outcome measure for the external test cohort was analogous to PROTECT, except the use of other 5-ASA therapy in addition to Pentasa was permitted.

### Image Pre-Processing

We implemented a two-step pre-processing strategy comprised of stain normalization and patch generation. We applied Python Staintools library^15^, a structure-preserving color package on the whole slide images (WSI), to standardize the slides with one WSI identified as the benchmark image. We first undertook brightness normalization using Luminosity Standardizer^16^, followed by stain normalization with the Vahadane method^16^. The digitally driven stain normalization process allows standardizing the stain color appearance of a source image with respect to a reference image (also referred to as the target image), with no specific, laboratory pre-analytical, procedures protocols, or specific expertise required (Supplementary Figure 1). We generated patches of 512*512 pixels and undertook experiments to determine the optimal overlap ratio and brightness threshold parameters. The overlap ratio indicates the overlap between patches, with the aim to provide sufficient coverage of the WSI, with the brightness threshold determining informative from non-informative patches. We applied the same imaging pre-processing parameters on both the PROTECT and Sickkids rectal biopsy slides.

### Histomic Features

Algorithms have been manually engineered, to extract distinctive characteristics and repeated patterns from histopathology images, that can be used as input features in machine learning models. These interpretable features have been referred to as histomics. The features capture the texture (spatial arrangement), morphology/shape, color, inter-voxel patterns, and orientations in a given image. We constructed five different classes of histomic features: nuclei, histogram-based, hue saturation value (HSV) color features and two texture features, gray level co-occurrence matrix features (GLCM) and local binary pattern (LBP) to capture information at the patch-level. For example, LBP feature creates a binary pattern, by comparing the intensity of each pixel in an image to the intensity of its neighboring pixel and encodes whether it is darker or brighter. To extract nuclei features we applied the Otsu threshold method, which can automatically separate an object of interest (e.g., nuclei), at a given threshold from the background tissue. Delaunay triangulations, and Voronoi diagram algorithms were applied to understand the spatial relationships between the nuclei. Feature computation details are described in the Supplementary Methods. Imaging data was read by SimpleITK package in Python^17^, and GLCM and LBP features generated by skimage packages^18^ (graycomatrix, local_binary_pattern). We utilized HistomicsTK, a Python package for the analysis of digital pathology images, to count the number of nuclei^19^. Features were implemented using self-developed functions without relying on pre-existing packages or libraries.

### Model Training and Identification of Optimal Features

We trained histomic features on 13 machine learning models (Naïve Bayes, Bagging, AdaBoost, CatBoost, and Tree-based) and logistic regression with 5-fold cross-validation, for patch-level classification. We fine-tuned the hyperparameters by grid search^20^ (Supplementary Figure 2). Models were implemented and built by Scikit-learn library^21^.

Feature importance was determined by the mean decrease in GINI (MDG), a measure of how each variable discriminates each image into their correct class, averaged across all decision trees^22, 23^. Features with higher MDG, have the greatest predictive power and are most important for classification^24^. The feature importance was computed using the Scikit-Learn ‘features_importance’ function and was normalized. We selected the optimal features for classification and re-trained the patch-level models. We undertook an alternative approach to further understand the impact of each feature class. We trained the 5-class features independently and determined the most discriminative features based on the MDG. We combined the optimal features into a single feature pool and re-trained the ML classifier. Slide-level prediction was defined by threshold voting. The voting thresholds were evaluated from 0.4 to 0.6, with CSFR ratio as a hyperparameter.

### Evaluating Prediction Model Performance

The performance of patch-level and whole slide image (WSI) models was evaluated using area under the receiver operating characteristic curve (AUROC), accuracy, precision, sensitivity, specificity, F1 score, and 95% confidence intervals obtained by 5-fold boot strapping. The F1 score combines precision and recall into one metric, and ranges from 0 to 1, 0 indicating poor model performance. We used the DeLong test to assess the performance difference between the AUROC of the various predictive models.

### Feature Interpretation

To evaluate the impact and importance of the optimal features on remission prediction, we generated the SHapley Additive explanation (SHAP) value^24^. The SHAP value measures the contribution of the feature value to the prediction value. Positive SHAP values (direction of x-axis) were indicative of CSFR, and higher mean feature value (in red color) a positive impact on prediction.

## RESULTS

### Cohort Characteristics

The training cohort consisted of 292 treatment naïve pediatric UC patients from the multicenter PROTECT study. The median age at presentation was 12.7 years (IQR 11, 15), 154 (53%) were male, 242 (83%) White and 17 (6%) Black. 118 (41%) patients achieved the primary outcome of corticosteroid free remission (CSFR) on mesalamine therapy alone at one year after diagnosis. Table 1 summarizes the demographic, phenotypic and histological features. The independent validation cohort consisted of 113 pediatric UC patients from a single center. The median age was 13 years (IQR 11,15), and 60 (53%) were male, 56 (51%) White, 23 (21%) South Asian and 8 (7%) Black. Forty percent achieved CSFR on 5-aminosalyciate therapy alone. The clinical disease severity at presentation differed between the two groups, medium PUCAI of PROTECT patients being 50 (IQR 35, 65) compared to 60 (IQR 40, 75) in the validation cohort, p=0.01 (Supplementary Table 1).

### Identification and Quantification of Histomic Features

Figure 1A depicts the pre-processing workflow to generate informative patches from our whole slide digitized H&E image (WSI). The figure outlines our two-step pre-processing strategy compromised of stain normalization and patch generation. We adopted a brightness ratio of 0.8 and overlap patch ratio of 0.25, generating 187571 informative 512*512 patches (Supplementary Figure 3) from the 292 WSI from the PROTECT data, and 85842 patches from the 113 WSI from the Sickkids data. These patches were used to compute the histomic features for model training. Histomic features are objectively quantifiable and interpretable, representing various morphological architectures within the tissue. We computed 250 histomic input features at the patch-level from five classes: 64 histogram-based features, 156 gray level co-occurrence matrix (GLCM), 10 local binary pattern (LBP), 9 hue saturation value (HSV) color features, and 11 nuclei structure features (Figure 1B).

**Figure 1:**
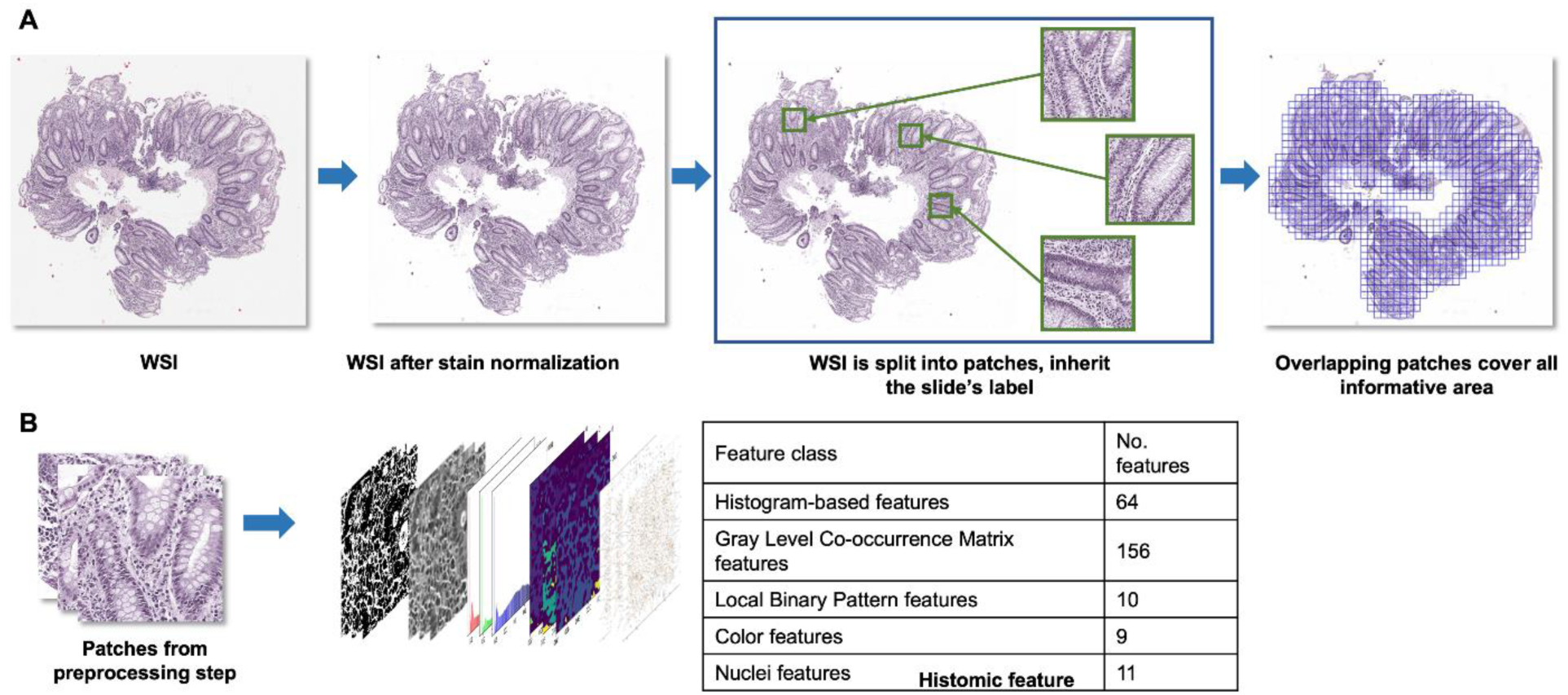
Histomic feature extraction approach. **(A)** A two-step pre-processing approach, including stain normalization and patch generation. The Vahadane method was applied to stain normalize, and two thresholds were used for patch generation process to select informative patches, and to provide sufficient coverage of the WSI (B) We then computed 250 histomic input features at the patch-level for model training, from five classes: Histogram-based features, color features, nuclei features and two texture features: Gray level co-occurrence matrix features, local binary pattern features

Histogram-based features represent the distribution of color or intensity values in an image using a histogram^25^. The mean and standard deviation of the pixel values in a histogram can be used to represent the brightness and contrast of an image, whilst the skewness and kurtosis describe the texture. GLCM texture features describe the spatial relationship between pixel intensities in an image^26^. LBP features compare the gray-level intensity of each pixel with its neighboring pixels^27^. Nuclei features were generated based on three different polygon methods (Otsu’s threshold, Delaunay triangulations, and Voronoi diagrams), and features were determined from the pixel value from each polygon. Five nuclei features used the Otsu’s algorithm which is a thresholding method to segment the images into small objects^28^ (Figure 2).

**Figure 2:**
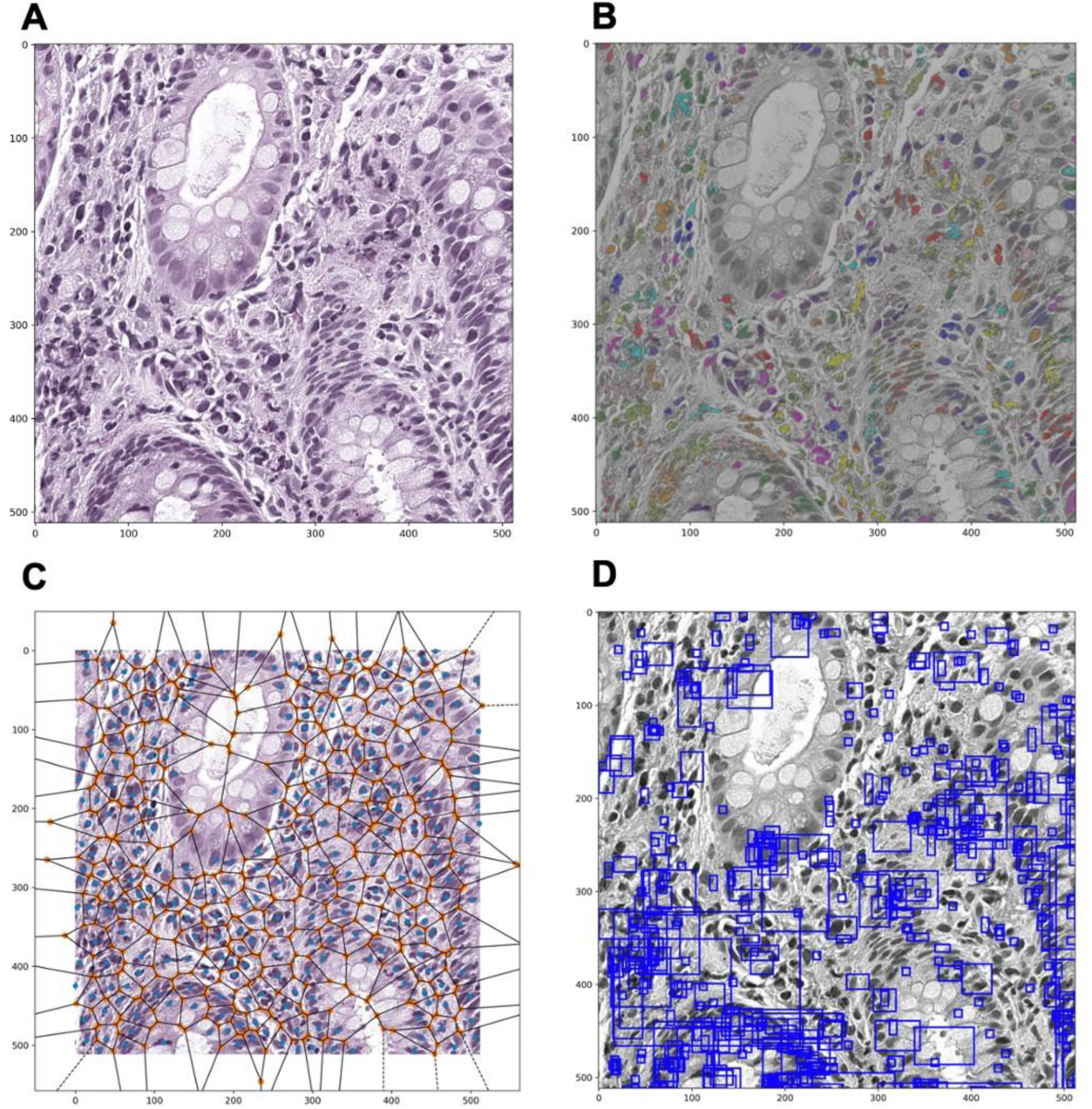
Example images of nuclei Histomic features. **(A)** Image shows hematoxylin and eosin after stain normalization, **(B)** an image of nuclei segmentation, **(C)** Otsu threshold polygon and **(D)** nuclei bounding boxes generated by histomics TK^19^

### Predictive Performance of the Patch-level Histomic Model

187571 informative patches from 292 patients were trained with 5-fold cross validation on 13 machine learning classifiers and logistic regression (Supplementary Table 2A). The optimal model trained on 250 histomic features at the patch-level was random forest (RF). The AUROC was 0.92 (95%CI: 0.89, 0.95) and accuracy 0.92 (95%CI: 0.90, 0.94), compared to logistic regression 0.52 (95%CI: 0.44, 0.60) and 0.53 (95%CI: 0.45,0.60), respectively (Figure 3). Eighteen optimal features were selected based on the ranking of the feature importance computed by the mean decrease in Gini (MDG) (Figure 3A, Supplementary Figure 4). These features consisted of three gray level co-occurrence matrix (GLCM), eight local binary patterns (LBP), two hue saturation value (HSV) and five nuclei features. The best model trained on the 18 features at the patch-level, was random forest model (RF), with an AUROC of 0.88 (95%CI: 0.85,0.92) and accuracy of 0.90 (95%CI: 0.80, 1.00) (Supplementary Table 2B). Collectively, these analyses identified a set of 18 pre-treatment rectal histomic features associated with steroid free clinical remission. The optimal performance characteristics were achieved by training the data at the patch-level on a random forest classifier.

**Figure 3:**
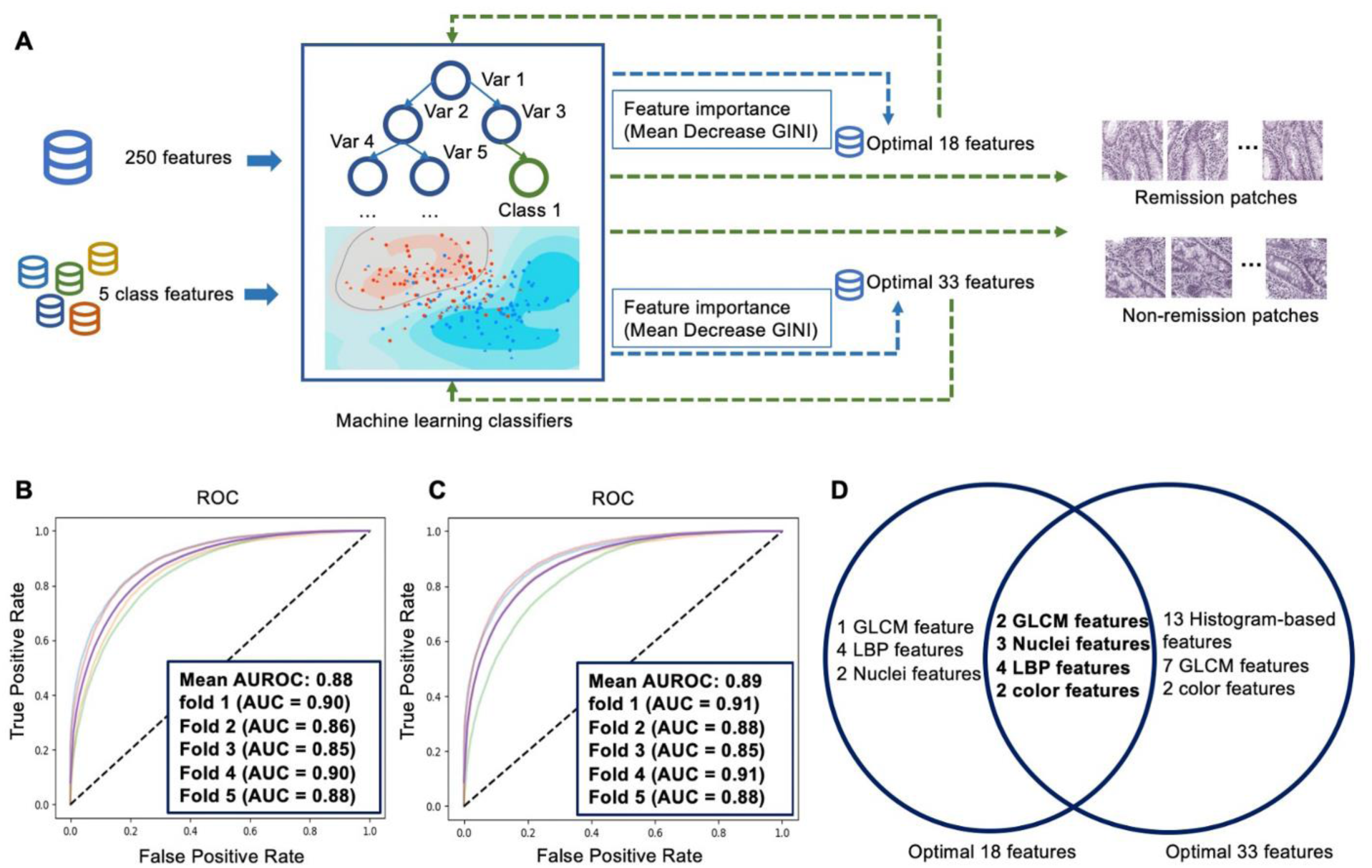
Overview machine learning approach and comparative patch-level predictive model performance. **(A)** Overview of histomic predictive machine learning approach, shows two parrallel approaches. Thirteen machine learning models trained using i) the entire 250 features and ii) 5 class features independently. Feature importance determined by mean decrease in GINI (MDG) for each approach and re-trained to classify corticosteroid free clinical remisson (CSFR) on mesalamine alone at one year. Plot **(B)** shows under the receiver operative curve (AUROC) with 95% confidence interval (CI), 5-fold cross validation (CV), for patch-level performance using the optimal 18 features derived from the 250 features. Plot **(C)** shows AUROC with 95% CI and 5-fold CV, for patch-level performance using the optimal 33 features from the 5-class feature approach. Venn diagram **(D)** shows shared histomic features between the 18 and 33 optimal features, and includes 12 features: GLCM_contrast_3_2, GLCM_contrast_1_2, LBP_2_5, LBP_2_0, LBP_2_7, LBP_2_1, LBP_2_6, H_mean, Otsu_equivalent_diameter, Otsu_area, Otsu_perimeter, Otsu_extent.

To evaluate the importance of each feature class, we also trained each class using the two top performing models (Supplementary Table 3A and 3B). Random forest outperformed extra trees at the patch-level for each class. The AUROC of histogram-based features (n=64) was 0.85 (95%CI: 0.82, 0.88), AUROC for GLCM (n=156) was 0.87 (95%CI: 0.84, 0.90), AUROC of LBP features (n=10) was 0.83 (95%CI: 0.80, 0.86), AUROC of color (n=9) features was 0.80 (95%CI: 0.78, 0.82), and AUROC of nuclei features (n=11) was 0.80 (95%CI: 0.76, 0.83). For each class, the optimal features based on MDG were selected, of 13 histogram-based features, 4 LBP features, 9 GLCM features, 4 color features, and 3 nuclei features. A total of 33 features were identified and were trained on RF and Extra Trees (ET) models. Random forest was the best model, with a patch-level AUROC of 0.89 (95%CI: 0.85, 0.93) and accuracy 0.90 (95%CI: 0.87, 0.92) (Supplement Table 2B).

### Predictive Performance of the Whole Slide Image Histomic Model

To distinguish CSFR from non-CSFR at the individual slide-level from patch-level predictions, we determined a voting threshold of 0.475 (optimal true positive and negative predictive value). Supplementary Figure 5 summarizes the experimental findings of various voting thresholds. The AUROC and accuracy at the WSI from the patch-level model using the entire set of 250 histomic features was 0.87 (95%CI: 0.73, 1.00) and 0.90 (95%CI: 0.80, 1.00) and with 33 features 0.89 (95%CI: 0.82, 0.94) and 0.90 (95%CI: 0.80, 1.00), respectively. The model trained using 18 optimal features was comparable with models trained on 250 features, with an AUROC of 0.89 (95%CI: 0.71,0.96) and accuracy of 0.90 (95%CI: 0.80, 1.00) (Table 2A). The DeLong test demonstrated no significant statistical difference between the predictive performance of the random forest model using 18 versus 33 features, p-value of 0.59.

**Table 2A:** Whole slide image model performance metrics using the 18 optimal Histomic features.

|  | **Sensitivity** | **Specificity** | **Precision** | **F1 score** | **Accuracy** | **AUROC** |
| --- | --- | --- | --- | --- | --- | --- |
| ***PROTECT cohort*** |  |  |  |  |  |  |
| Random Forest | 0.84<br>(0.78,0.90) | 0.94<br>(0.82,1.00) | 0.91<br>(0.82,1.00) | 0.87<br>(0.82,0.92) | 0.90<br>(0.80,1.00) | 0.89<br>(0.71,0.96) |
| Logistic regression | 0.47<br>(0.30,0.63) | 0.50<br>(0.41,0.58) | 0.59<br>(0.50,0.67) | 0.48<br>(0.40,0.56) | 0.48<br>(0.42,0.54) | 0.48<br>(0.41,0.56) |
| ***SICKKIDS cohort*** |  |  |  |  |  |  |
| Random Forest | 0.78<br>(0.49,1.00) | 0.91<br>(0.81,1.00) | 0.84<br>(0.57,1.00) | 0.82<br>(0.59,1.00) | 0.85<br>(0.63,1.00) | 0.85<br>(0.75, 0.95) |
| Logistic regression | 0.42<br>(0.34,0.50) | 0.43<br>(0.39,0.49) | 0.53<br>(0.50,0.59) | 0.43<br>(0.40,0.47) | 0.48<br>(0.44,0.51) | 0.47<br>(0.40,0.55) |

**Table 2B:**
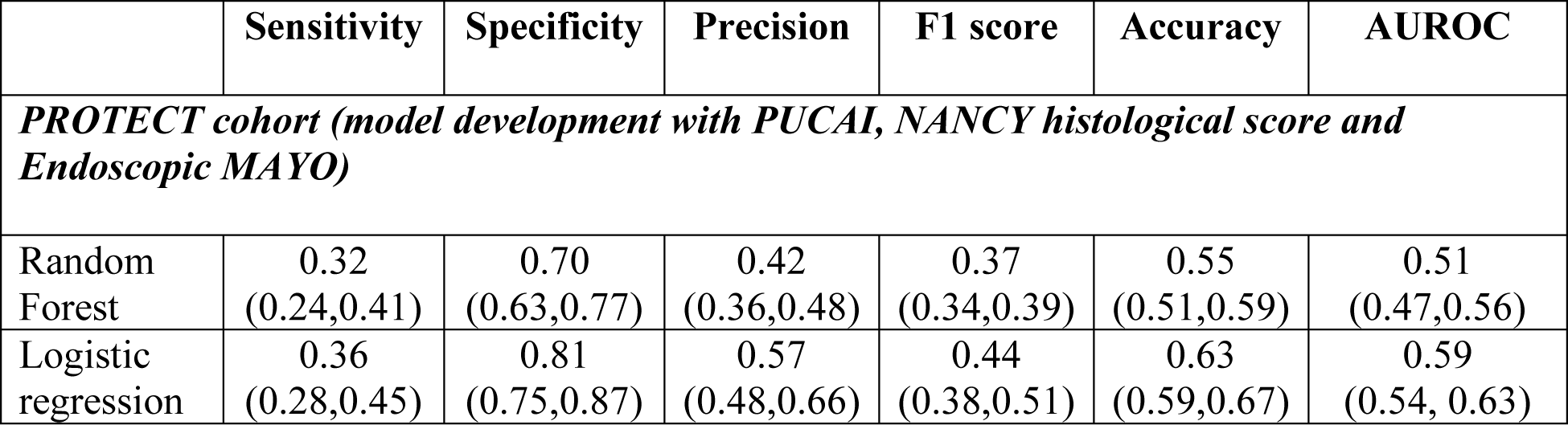
Model performance metrics using baseline clinical disease severity variables.

### Feature Validation on an External Cohort

We evaluated the histomics feature set on a real-world cohort of pediatric UC patients. We applied the optimal 18 histomic features on an independent external cohort. The AUROC and accuracy at the patch level was 0.88 (95%CI: 0.84, 0.91) and 0.88 (95%CI: 0.82, 0.92) respectively. Similarly, at the WSI level AUROC of 0.85 (95%CI: 0.75, 0.95), and accuracy of 0.85 (95%CI: 0.75, 0.95) respectively (Table 2A). The optimal histomics feature in an external validation cohort exhibited comparable performance (Table 2A).

### Interpretability of Histomic Features

The interpretability of the optimal features was evaluated by the Shapley Additive exPlanations (SHAP) value, as illustrated in Figure 4A. The y-axis indicates the feature. The x-axis indicates the SHAP-value, a positive value indicative of CSFR and a negative value of non-CSFR. The magnitude of the feature value is represented by a color bar, red being high and blue low. Each individual patient is represented by a dot. We noted nuclei features with a higher value, Otsu_equivalent_diameter, Otsu_area, and Otsu_perimeter, have a positive impact upon the likelihood of CSFR. Conversely, a high value of LBP_2_5 has a negative impact (negative SHAP value) on the likelihood of CSFR. Figure 4B visualizes the feature Otsu_equivalent_diameter at the slide level. We observed a similar trend in our external cohort (Supplementary Figure 6).

**Figure 4:**
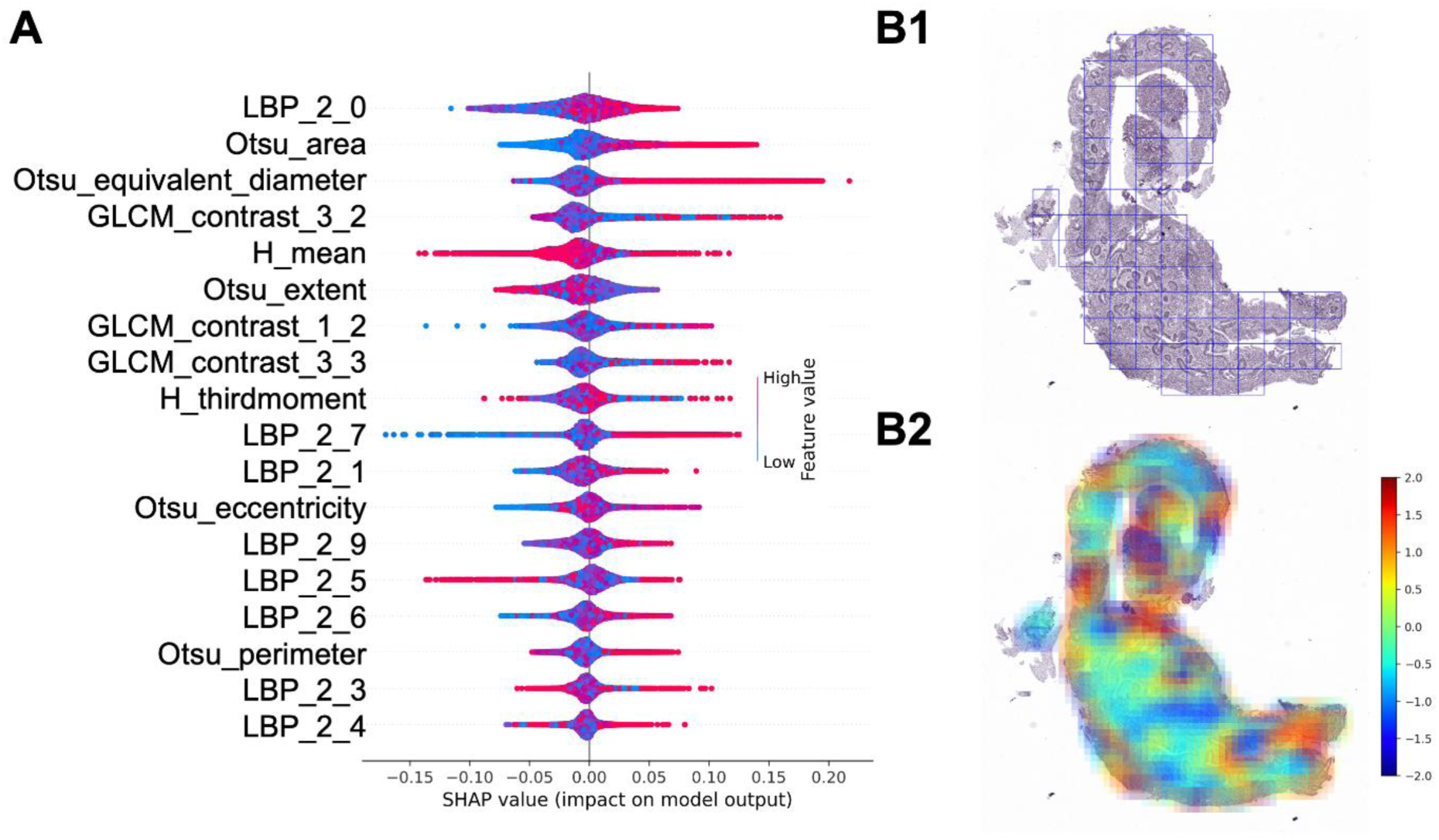
Histomic feature importance represented by the SHapley Additive exPlanations (SHAP) values. Plot **(A)** shows the relationship between the 18 histomic features and the outcome of CSFR with mesalamine alone at one year. Positive SHAP-values (x-axis) are indicative of clinical remission, and negative value of non-remission. The magnitude of each feature value is represented by the color bar, red being high and blue low. Features include LBP: Local Binary Pattern, GLCM: Gray Level Co-occurrence Matrix; Histogram features: H_third moment and H_mean and Nuclei features: Otsu_perimeter, Otsu_eccentricity, Otsu_area and Otsu_equivalent_diameter. Image **(B1)** shows non-overlapping patches of H&E stain normalized whole slide images, and **(B2)** heatmap of the Otsu equivalent diameter, a nuclei feature and the color bar representing feature values, high value in red/brown, low value in blue.

## DISCUSSION

In this multicenter study, we discovered and externally validated 18 histomic features extractable from routine clinical rectal histopathology samples collected at diagnosis. These features accurately predicted which pediatric UC patients would achieve CSFR on mesalamine therapy alone.

In the PROTECT inception cohort study of standardized corticosteroid and mesalamine induction therapy, we evaluated features at diagnosis and following four weeks of induction therapy predictive of week 52 CSFR on mesalamine alone. We found patients with a pediatric ulcerative colitis index <45 and hemoglobin ≥10g/dL at diagnosis, together with achieving clinical remission by four weeks following diagnosis, were more likely to experience this outcome.^12^ The clinical and laboratory features demonstrated only modest predictive utility in classifying patients for this outcome, which is in contrast to the 18 histomic features, exhibiting superior predictive capability.

Our analysis of routine histopathology demonstrated children presenting with more severe clinical disease activity at diagnosis exhibited lower levels of rectal eosinophilic inflammation, and higher levels of surface epithelial villiform changes.^2^ Consistent with this, patients with a higher rectal biopsy eosinophil count were less likely to escalate to anti-TNF therapy.^1, 12^ However, no standard histopathology features were found to be predictive of the primary outcome, CSFR on mesalamine alone. A recent study utilized digitized colonic biopsies from 273 adult UC patients to automate classification of histologic severity, and to predict future flares of disease.^29^ The model accurately distinguished colonic samples with histopathologic activity from those in histologic remission, and performed similarly to expert pathologists in predicting flares of clinical disease activity. Similarly, a computer-aided diagnosis system automated the PICaSSO Histologic Remission Index (PHRI), a simplified scoring system that detects mucosal neutrophils, reliably identifying active from quiescent UC in a subset of 138 biopsies.^30^ Consistent with this, in the current study we have discovered and validated 18 predictive rectal histomic features extractable from routine clinical histopathology specimens. Importantly, the machine-learning based histomic features models were superior to logistic regression, and models based on clinical, endoscopic, and histopathologic measures of severity for patient classification. These recent studies emphasize the likely high impact of incorporating automated histopathology analysis into UC patient classification, ideally with further refinement of these approaches in the context of additional cohort studies and clinical trials.

Our study has several strengths, but also some limitations. We applied a novel machine-learning pipeline to derive and test histomic features associated with an important clinical outcome in pediatric UC patients. Our discovery cohort was from a large multicenter study which utilized standardized corticosteroid and mesalamine induction therapy, while our validation cohort reflected real world experience. Despite increased clinical and endoscopic disease severity in the validation cohort, the frequency of steroid-free remission with mesalamine alone was comparable to the discovery cohort. Moreover, the 18 optimal rectal histomic features exhibited similar test characteristics for patient classification in the two cohorts. This may suggest shared biology underlying histomic features and treatment responses, not accounted for by current clinical severity measures. Following further validation in adult UC patients, these histomic features are likely to have strong generalizability. Moreover, following digitization and pre-processing, the rectal biopsy slides obtained for routine clinical care yielded reliable histomic data, supporting widespread testing of this approach to supplement routine histopathologic assessments. Also, histomic features are agnostic and unlike deep learning are interpretable, reproducible, and easily employed in digital pathology for automation.

In the current study, we have validated 18 rectal histomic features which when incorporated in a machine learning model predicted steroid-free remission on mesalamine alone in children with Ulcerative Colitis. These morphometric features capture the properties within the tissue, and further support the development of an agnostic automated histopathology based predictive tool for classifying treatment response in UC patients.

## Abbreviations

UC: Ulcerative colitis
CSFR: Corticosteroid-free remission
WSI: Whole slide images
ML: Machine learning
H+E: Hematoxylin and eosin
PUCAI: Pediatric ulcerative colitis activity index
HSV: Hue saturation value
GLCM: Gray level co-occurrence matrix
LBP: Local binary pattern
MDG: Mean decrease in GINI
AUROC: Area under the receiver operating characteristic curve
SHAP: Shapley Additive explanation
RF: Random forest
ET: Extra trees

## Data Availability

All data produced in the present study are available upon reasonable request to the authors

https://repository.niddk.nih.gov/

