## Supplementary file for "Machine Learning-Based Prediction of Pediatric Ulcerative Colitis Treatment Response using Diagnostic Histopathology"

#### Supplemental Methods: Texture feature Definitions

**Histogram-based features**

In this study, we extracted histogram-based features from the histogram of pixels per patch using the methodology described in the main methods section. Histogram features are the most fundamental features extracted from imaging data. We generated a histogram for each patch using the aforementioned methodology to extract these features. This histogram represents the distribution of pixels in the image, with each grayscale value represented as a single integer from 0 to 255 (black to white, respectively 0 to 1). The histogram plot then counts the number of pixels in a specific pixel range. Based on this information, we generated 16 statistical histogram features, including minimum, maximum, mean, median, variance, energy, entropy, ten pentiles, ninety percentile, interquartile range, range, mean absolute deviation, robust mean absolute deviation, root mean square error, skewness, and kurtosis. These lower-order statistical features accurately describe the texture of each patch. Overall, we generated a total of 64 histogram-based features.

**GLCM-based features**

To generate the gray level co-occurrence matrix, one pixel was labeled as the reference, and the neighboring pixels were labeled as neighbors. Each patch may contain several pairs, consisting of one reference pixel and one neighbor pixel with different directions and displacements. As a result, the gray level co-occurrence matrix consists of the count of each pair of pixels, with the elements representing the number of pixel occurrences with gray levels $\boldsymbol{i}.\ \boldsymbol{j}$, represented by a displacement of one pixel in the direction of zero degrees. In our study, we defined the co-occurrence matrix using four directions (0°, 45°, 90°, and 135°) and three displacement vectors (1px, 3px, 5px) as the length between the reference pixel and neighbor pixels. We extracted a variety of features, including energy, contrast, correlation, variance, GLCM inverse difference of moment, GLCM sum average, GLCM sum entropy, sum, variance, GLCM entropy, GLCM difference variance, and GLCM difference entropy. Overall, we generated a total of 156 GLCM-based features, which are expected to provide valuable insights into our study's imaging data.

$$contrast = \sum_{\boldsymbol{i,j}} |\boldsymbol{i}-\boldsymbol{j}|^2 \boldsymbol{y}_{\boldsymbol{i,j}} \tag{1}$$

$$correlation = \sum_{i,j} \frac{(i-\mu_i)(j-\mu_j)p_{i,j}}{\sigma_i \sigma_j} \tag{2}$$

$$energy = \sum_{i,j} p_{i,j}^2 \tag{3}$$

$$entropy = -\sum_{i,j} p_{i,j} log_2 p_{i,j} \tag{4}$$

**LBP-based features**

In our study, we used each pixel as the reference, and the pixels around that reference were considered neighbor pixels (with a displacement of 2). For instance, if we randomly select a pixel, we compare its intensity with all nine neighbor pixels. If a neighbor pixel is larger than the central pixel, then the binary pattern is assigned a value of 1; otherwise, the binary pattern is assigned a value of 0. Once all binary patterns are computed, we apply a weighted matrix consisting of values

1, 2, 4, 8, 16, 32, 64, and 128 in a circular manner on the binary pattern matrix. In our study, we used radius values of 2 and 8 around points for LBP features. Based on the LBP values, we extracted 10 histogram bins' values of LBP, which served as the LBP features in our study.

**Color-based features**

We utilized HSV (Hue, saturation, and value or brightness) color space to create suitable features in medical image preprocessing and classification to help researchers find more helpful content. Converting patches into HSV space, we compute one-stage moment (mean value for H, S, and V), secondary moments (standard deviation for H, S, and V), and three order moment variances for H, S, and V. Therefore, our study covered 9 HSV color features.

$$the\ mean = \sum_{i=1}^{n}\sum_{j=1}^{m}\frac{x_{ij}}{mn} \tag{5}$$

$$variance = \frac{1}{mn}\sum_{i=1}^{n}\sum_{j=1}^{m}\left(X_{ij} - mean\right)^2 \tag{6}$$

$$stddev = \sqrt{variance}, where\ X_{ij}\ is\ the\ pixel\ value\ of\ i^{th}\ row\ and\ j^{th}\ column \tag{7}$$

**Nuclei features**

Our study utilizes HistomicsTK to count the number of nuclei. HistomicsTK provides a pipeline for nuclei segmentation from color normalization, color deconvolution, and segment nuclei. In the color normalization process, utilizing the mean and standard deviation of the reference image in lab space, then perform Reinhard color normalization. The color map consists of hematoxylin, eosin, dab, and null (input contains only two stains) for the color deconvolution process. And then perform the standard color deconvolution. Several parameters are defined during this process for the segment nuclei process: minimum radius, maximum radius, local maximum search radius (which detects and segments nuclei using local maximum clustering), and minimum nucleus area (which is used to filter the small objects). We defined those parameters with guidance from the pathologist from Cincinnati Children’s Hospital. The Otsu threshold generates the second nuclei feature in order to determine the small nuclei regions; we computed average Otsu area, average Otsu perimeter, average Otsu extent, average Otsu equivalent diameter, and Otsu eccentricity. We also utilized Delaunay triangulations and Voronoi diagrams to create the polygon based on patches and computed the average area, and the site disorder. The average area per patch is computed by equation 8

$$\widehat{\mathcal{N}} = \frac{1}{m}\sum_{j=0}^{m}\mathcal{N}^{A}(p_j) \tag{8}$$

$$\widehat{D} = 1/(1 + \frac{\sigma^A}{\widehat{\mathcal{N}^A}}) \tag{9}$$

where m is the number of polygons created by the spatial graph, P is the polygon, and $\mathcal{N}^{A}(p_j)$ is the area of number j polygon. Therefore, the disorder of the polygon area is computed by * where $\sigma^A$ is the standard deviation of $\mathcal{N}^{A}(p_j)$. In total, there are 11 features from nuclei-based features.

Supplementary table 1: demographic, clinical, and histological characteristics comparison of internal and external Cohorts

| | **PROTECT Internal** | **TORONTO/ SICKKIDS External** | **p-value** |
|---|---|---|---|
| **Age years** | 12.7 (11,15) | 13 (11, 15) | 0.56 |
| **Male** | 154 (53%) | 60 (53%) | 0.95 |
| **Race** | | | |
| **Black/African American** | 17 (6%) | 8 (7%) | |
| **White/Caucasian** | 242 (83%) | 56 (51%) | |
| **South Asian** | - | 23 (21%) | 0.05 |
| **East Asian** | 12 (4%) | 3 (3%) | |
| **Mixed** | 14 (5%) | 20 (18%) | |
| **Endoscopic Mayo** | | | |
| **Mayo 1** | 38 (13%) | 13 (11%) | |
| **Mayo 2** | 158 (54%) | 48 (43%) | 0.05 |
| **Mayo 3** | 96 (33%) | 52 (46%) | |
| **PUCAI** | 50 (35, 65) | 60 (40, 75) | 0.01 |

SUPPLEMENTARY FIGURE1: STAIN NORMALIZATION

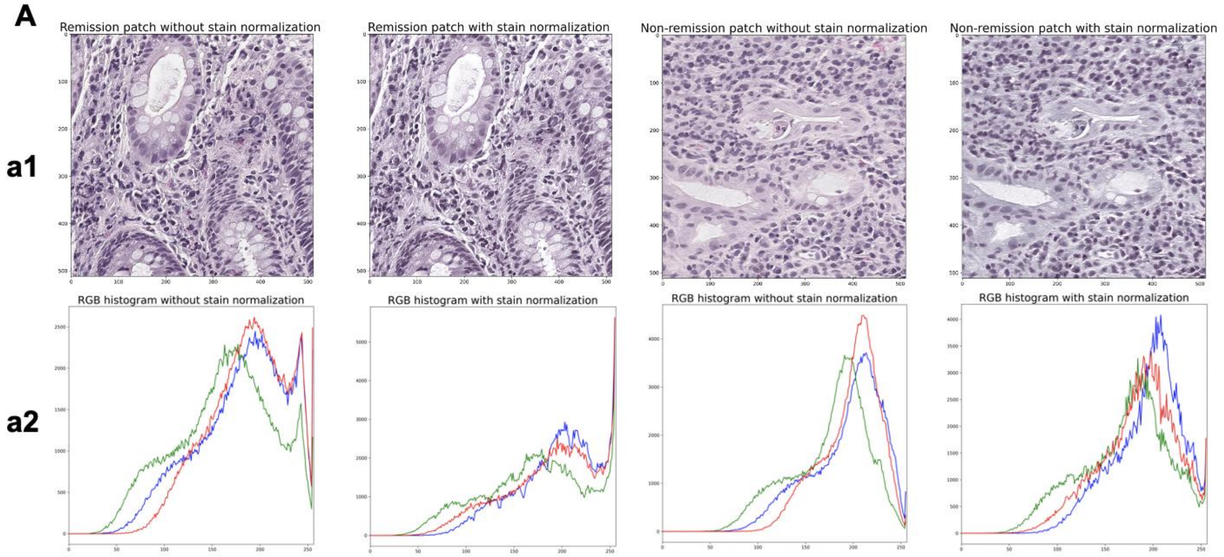

The benchmark WSI was 88550 as slide_id. Before applying the slide-level normalization and patches generation, we loaded svs format slides into QuPath, manually cropped them into smaller sizes, and stored them into original png format slides to reduce the computation time once we started generating patches because WSIs contained a large area of white space without information area. The improvement of the Vahadane method was that this method did not change the original tissue compared with the color normalization and also normalized the image at patch-level instead of slide-level directly. A1 shows patches with and without stain normalization. A2 shows the histogram of patches with and without stain normalization.

### SUPPLEMENTARY FIGURE 2: GRID-SEARCH FOR TREE-BASED MODEL

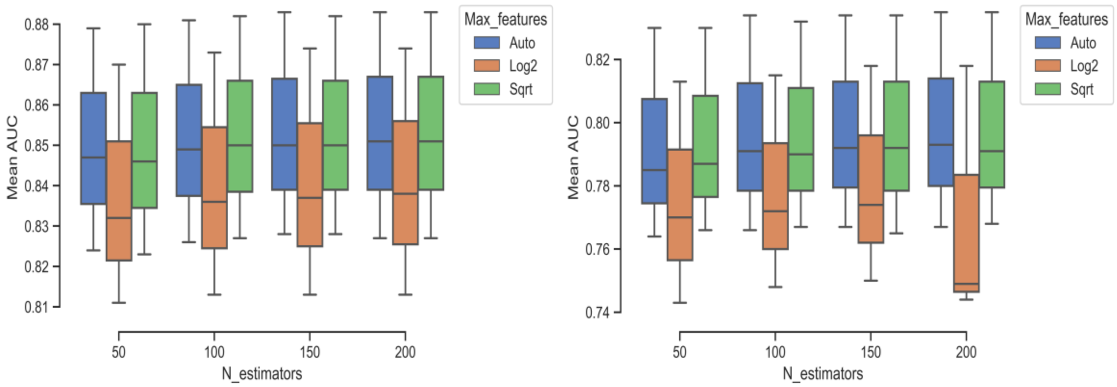

For the tree-based models, we defined the number of trees from 50 trees to 200 trees with 50 intervals, the number of features for each split by both 'auto', 'sqrt', and 'log2' methods, and the minimum number of samples for each split from 100 to 300 samples with 100 as the interval. The features were sorted by the GINI index, and the model was trained with bootstrap.

The best performance of the extra tree model is defined by: 100 as min split samples, 200 as number of trees, and uses the square root of the entire number of features as the maximum number of features. The optimal performance of the random forest model is defined by: 100 as min split samples, 150 as number of trees, and using auto as the maximum number of features. With those parameter settings, random forest model performed better than extra trees model on the patch-level prediction.

#### SUPPLEMENTARY FIGURE 3: INFORMATIVE VS. NON-INFORMATIVE PATCHES

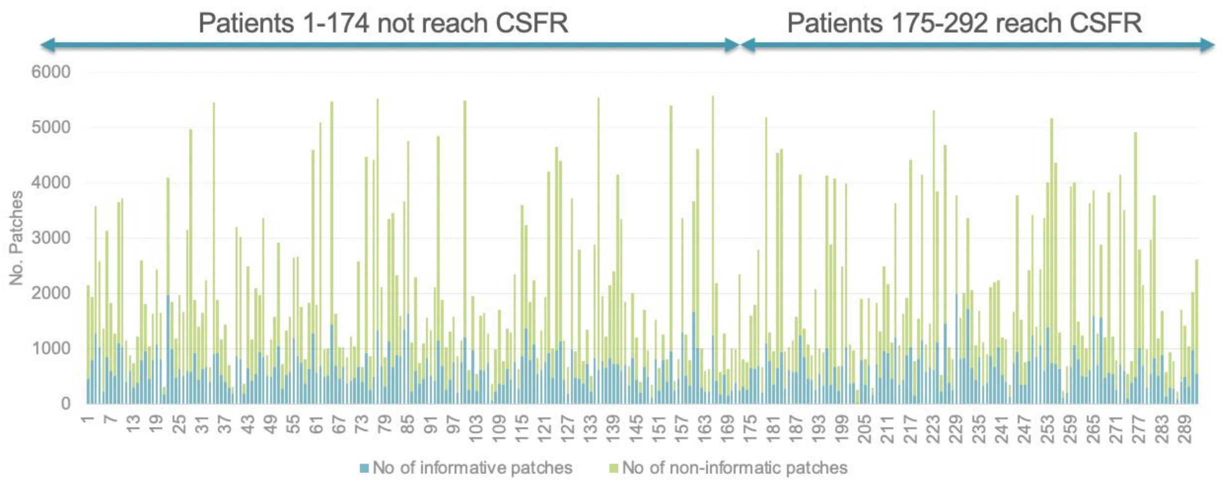

The overlap ratio represents the overlap between patches, and the brightness threshold helps with measuring the informatic or non-informatic patches, the ratio from 1.0 (which is non-overlap in x-coordinate), 0.5 (which has 256-pixel non-overlap in x-coordinate), 0.25 (which has 128-pixel nonoverlap in x-coordinate), and 0.125 (which has 64 pixel non overlap in x-coordinate). This paper uses 0.25 (overlap ratio) and 0.8 (brightness) as the selection thresholds to select the patches with information. The average number of informative patches per remission slide is 648, and the average number of informative patches per non-remission slide is 639. In our experiment, we identified information-containing patches using 0.25 (overlap ratio) and 0.80 (brightness) as the selection criteria.

#### SUPPLEMENTARY TABLE 2A: 13 MACHINE LEARNING MODEL PERFORMANCE (PATCH-LEVEL) AND LOGISTIC REGRESSION

| | **Sensitivity** | **Specificity** | **Precision** | **F1-score** | **Accuracy** | **AUC** |
|---|---|---|---|---|---|---|
| Random Forest | 0.95 (0.93, 0.97) | 0.89 (0.82, 0.97) | 0.93 (0.88, 0.98) | 0.93 (0.86, 0.95) | 0.92 (0.9, 0.94) | 0.92 (0.89, 0.95) |
| Extra Tree | 0.88 (0.84, 0.93) | 0.77 (0.65, 0.9) | 0.85 (0.74, 0.96) | 0.83 (0.86, 0.95) | 0.84 (0.81,0.87) | 0.83 (0.78, 0.88) |
| Logistic regression | 0.58 (0.43, 0.74) | 0.46 (0.42, 0.5) | 0.61 (0.52, 0.71) | 0.55 (0.31, 0.56) | 0.53 (0.45, 0.6) | 0.52 (0.44, 0.60) |
| Adaboost | 0.96 (0.92, 1.00) | 0.06 (0, 0.14) | 0.60 (0.45, 0.76) | 0.0 (0, 0.23) | 0.59 (0.45, 0.74) | 0.51 (0.49, 0.53) |
| BaggingClassifier | 0.77 (0.68, 0.85) | 0.27 (0.23, 0.32) | 0.61 (0.48, 0.74) | 0.38 (0.27, 0.39) | 0.56 (0.52, 0.61) | 0.52 (0.48, 0.56) |
| BernoulliNB | 0.52 (0.36, 0.67) | 0.46 (0.33, 0.58) | 0.59 (0.43, 0.74) | 0.42 (0.37, 0.44) | 0.48 (0.43, 0.54) | 0.49 (0.44, 0.53) |
| DecisionTreeClassifier | 0.60 (0.55, 0.66) | 0.42 (0.39, 0.46) | 0.61 (0.47, 0.74) | 0.48 (0.34, 0.49) | 0.53 (0.51, 0.54) | 0.51 (0.49, 0.54) |
| GaussianNB | 0.39 (0, 0.88) | 0.54 (0.14, 0.95) | 0.53 (0.43, 0.64) | 0.34 (0.3, 0.5) | 0.42 (0.31, 0.53) | 0.47 (0.41, 0.53) |
| Perceptron | 0.61 (0.38, 0.84) | 0.40 (0.33, 0.47) | 0.60 (0.51, 0.69) | 0.52 (0.27, 0.53) | 0.52 (0.41, 0.63) | 0.51 (0.4, 0.61) |
| SGDClassifier | 0.77 (0.56, 0.98) | 0.29 (0.16, 0.42) | 0.61 (0.48, 0.75) | 0.30 (0.29, 0.38) | 0.56 (0.49, 0.64) | 0.53 (0.48, 0.58) |
| RidgeClassifierCV | 0.75 (0.59, 0.92) | 0.28 (0.21, 0.35) | 0.61 (0.49, 0.73) | 0.35 (0.31, 0.35) | 0.56 (0.52, 0.59) | 0.52 (0.47, 0.57) |
| PassiveAggressive | 0.69 (0.61, 0.76) | 0.41 (0.25, 0.57) | 0.63 (0.46, 0.81) | 0.37 (0.34, 0.51) | 0.57 (0.49, 0.64) | 0.55 (0.5, 0.6) |
| GradientBoosting | 0.88 (0.79, 0.96) | 0.17 (0.14, 0.2) | 0.52 (0.48, 0.74) | 0.27 (0.21, 0.29) | 0.59 (0.52, 0.66) | 0.52(0.49, 0.56) |
| BaggingClassifier | 0.76 (0.68, 0.85) | 0.27 (0.23, 0.32) | 0.61 (0.48, 0.74) | 0.38 (0.27, 0.39) | 0.56 (0.52, 0.61) | 0.52 (0.48, 0.56) |

#### SUPPLEMENTARY TABLE 2B: PATCH-LEVEL MODEL PERFORMANCE ON THE THREE DIFFERENT FEATURE SETS

| | **Sensitivity** | **Specificity** | **Precision** | **F1-score** | **Accuracy** | **AUROC** |
|---|---|---|---|---|---|---|
| **250 HISTOMIC FEATURES** | | | | | | |
| Random Forest | 0.89 (0.82, 0.97) | 0.95 (0.93, 0.97) | 0.93 (0.88, 0.98) | 0.93 (0.86, 0.95) | 0.92 (0.9, 0.94) | 0.92 (0.89, 0.95) |
| Extra Tree | 0.77 (0.65, 0.9) | 0.88 (0.84, 0.93) | 0.85 (0.74, 0.96) | 0.83 (0.86, 0.95) | 0.84 (0.81,0.87) | 0.83 (0.78, 0.88) |
| Logistic regression | 0.46 (0.42, 0.5) | 0.58 (0.43, 0.74) | 0.61 (0.52, 0.71) | 0.55 (0.31, 0.56) | 0.53 (0.45, 0.6) | 0.52 (0.44, 0.6) |
| **18 OPTIMAL HISTOMIC FEATURES** | | | | | | |
| Random Forest | 0.85 (0.78, 0.93) | 0.91 (0.87, 0.95) | 0.90 (0.85, 0.96) | 0.90 (0.8, 0.91) | 0.89 (0.86, 0.91) | 0.88 (0.85, 0.92) |
| Extra Tree | 0.73 (0.62, 0.85) | 0.87 (0.81, 0.93) | 0.83 (0.74, 0.92) | 0.82 (0.67, 0.84) | 0.82 (0.78, 0.85) | 0.80 (0.75, 0.85) |
| Logistic regression | 0.44 (0.33, 0.55) | 0.57 (0.44, 0.69) | 0.60 (0.49, 0.71) | 0.52 (0.29, 0.54) | 0.51 (0.47, 0.55) | 0.50 (0.45, 0.56) |
| **33 OPTIMAL HISTOMIC FEATURES BASED ON 5-CLASS MODEL** | | | | | | |
| Random Forest | 0.86 (0.78, 0.94) | 0.92 (0.9, 0.95) | 0.91 (0.85, 0.96) | 0.91 (0.81, 0.92) | 0.90 (0.87, 0.92) | 0.89 (0.85, 0.93) |
| Extra Tree | 0.75 (0.64, 0.86) | 0.87 (0.82, 0.92) | 0.84 (0.74, 0.93) | 0.82 (0.68, 0.85) | 0.82 (0.78, 0.86) | 0.81 (0.76, 0.86) |
| Logistic regression | 0.47 (0.4, 0.54) | 0.56 (0.45, 0.67) | 0.61 (0.5, 0.72) | 0.53 (0.33, 0.54) | 0.52 (0.48, 0.56) | 0.51 (0.47, 0.56) |

#### SUPPLEMENTARY FIGURE 4A: FEATURE IMPORTANCE PLOT OF THE 250 FEATURES

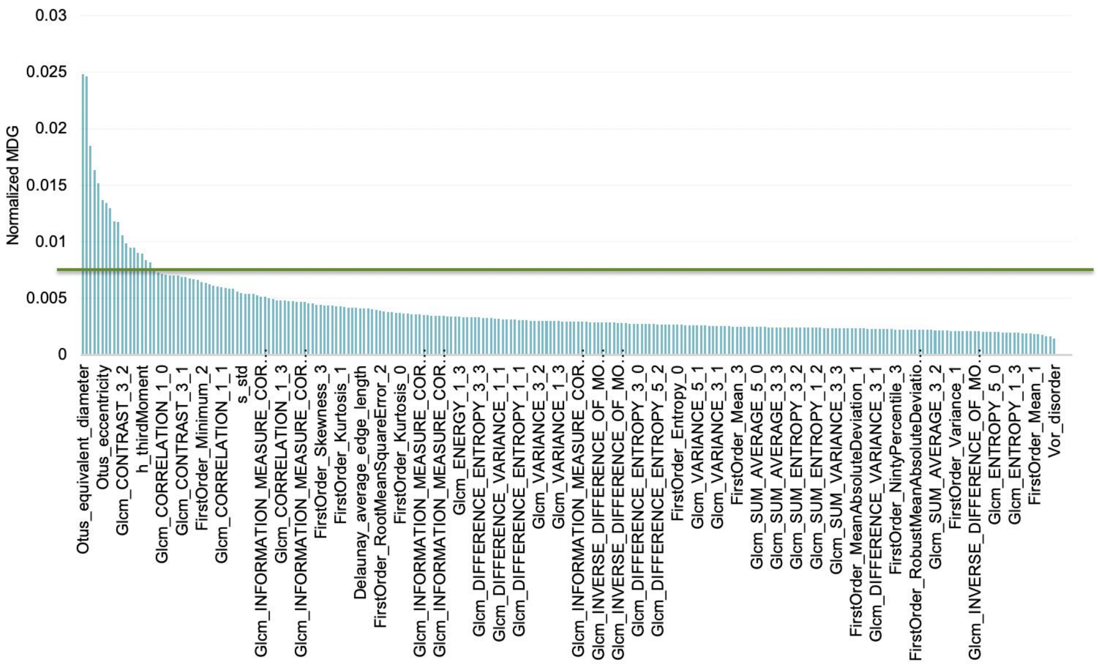

250 features importance sorted by Mean decrease in gini (MDG).

SUPPLEMENTARY FIGURE 4B: OPTIMAL 18 AND OPTIMAL 33 FEATURES WERE SELECTED FROM 250 FEATURES BY MDG

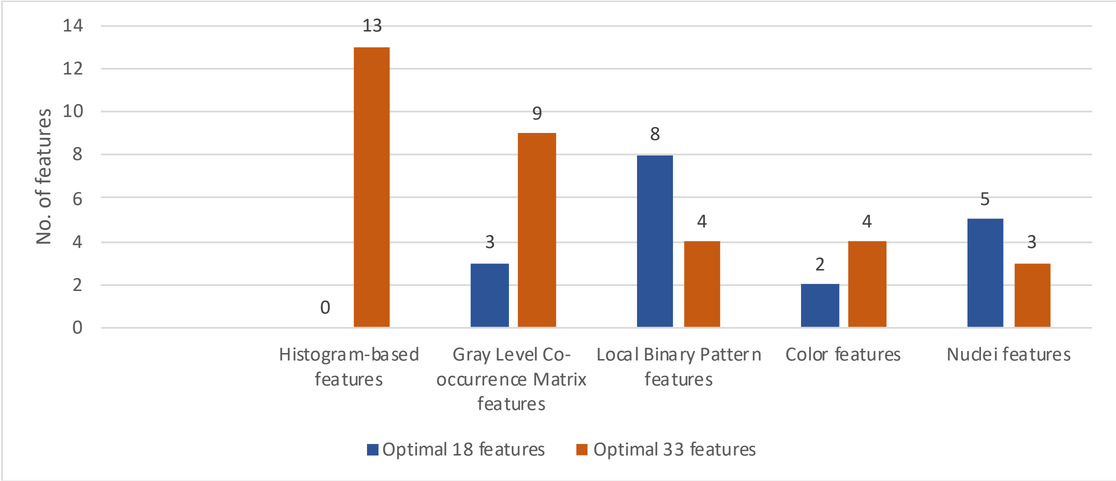

Optimal 18 features (A): features selected by feature importance from 250 feature set by cut-off value (0 histogram-based features, 3 gray level co-occurrence matrix features, 8 local binary pattern features, 2 color features, and 5 nuclei features)
Optimal 33 features (B): features selected by feature importance from each feature class set by cut-off value (13 histogram-based features, 9 gray level co-occurrence matrix features, 4 local binary pattern features, 4 color features, and 3 nuclei features)

#### Supplementary table 3: model performance on 5 class independently (patch-level)

| | **Sensitivity** | **Specificity** | **Precision** | **F1-score** | **Accuracy** | **AUC** |
|---|---|---|---|---|---|---|
| Histogram | 0.88 (0.84, 0.91) | 0.82 (0.75, 0.88) | 0.87 (0.79, 0.96) | 0.86 (0.75, 0.87) | 0.85 (0.83, 0.87) | 0.85 (0.82, 0.88) |
| GLCM | 0.91 (0.87, 0.94) | 0.84 (0.77, 0.91) | 0.89 (0.81, 0.97) | 0.88 (0.79, 0.89) | 0.88 (0.86, 0.90) | 0.87 (0.84, 0.90) |
| LBP | 0.86 (0.80, 0.91) | 0.80 (0.76, 0.85) | 0.87 (0.81, 0.92) | 0.86 (0.72, 0.87) | 0.84 (0.980, 0.86) | 0.83 (0.80, 0.86) |
| Color | 0.85 (0.81, 0.88) | 0.75 (0.69, 0.82) | 0.83 (0.74, 0.92) | 0.81 (0.69, 0.82) | 0.81 (0.80, 0.81) | 0.80 (0.78, 0.82) |
| Nuclei | 0.87 (0.82, 0.91) | 0.72 (0.65, 0.79) | 0.82 (0.73, 0.91) | 0.81 (0.68, 0.82) | 0.81 (0.80, 0.82) | 0.80 (0.76, 0.83) |

Supplementary Table 3A. Random forest on patch level performance

| | **Sensitivity** | **Specificity** | **Precision** | **F1-score** | **Accuracy** | **AUC** |
|---|---|---|---|---|---|---|
| Histogram | 0.82 (0.76, 0.88) | 0.72 (0.63, 0.82) | 0.81 (0.7, 0.92) | 0.78 (0.62, 0.81) | 0.78 (0.74, 0.82) | 0.77 (0.72, 0.82) |
| GLCM | 0.84 (0.79, 0.9) | 0.73 (0.62, 0.85) | 0.82 (0.71, 0.94) | 0.79 (0.66, 0.82) | 0.80 (0.77, 0.82) | 0.79 (0.74, 0.83) |
| LBP | 0.79 (0.71, 0.87) | 0.73 (0.67, 0.79) | 0.81 (0.74, 0.88) | 0.80 (0.6, 0.81) | 0.77 (0.72, 0.81) | 0.76 (0.71, 0.81) |
| Color | 0.77 (0.71, 0.83) | 0.65 (0.57, 0.73) | 0.77 (0.65, 0.88) | 0.72 (0.56, 0.74) | 0.72 (0.71, 0.73) | 0.71 (0.68, 0.75) |
| Nuclei | 0.82 (0.76, 0.88) | 0.59 (0.46, 0.71) | 0.74 (0.62, 0.87) | 0.69 (0.52, 0.72) | 0.72 (0.69, 0.75) | 0.70 (0.65, 0.76) |

Supplementary Table 3B. Extra tree classifier on patch level performance

#### SUPPLEMENTARY FIGURE 5: VOTING THRESHOLD

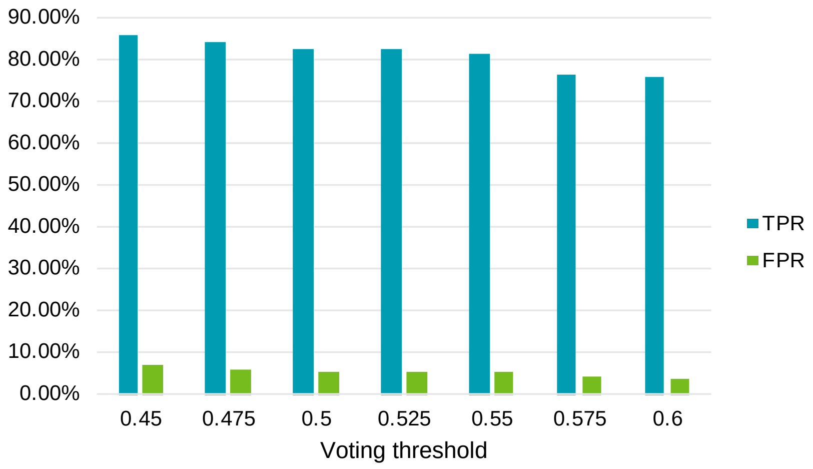

SUPPLEMENTARY FIGURE 5A. RANDOM FOREST SLIDE-LEVEL PERFORMANCE BY VOTING THRESHOLD

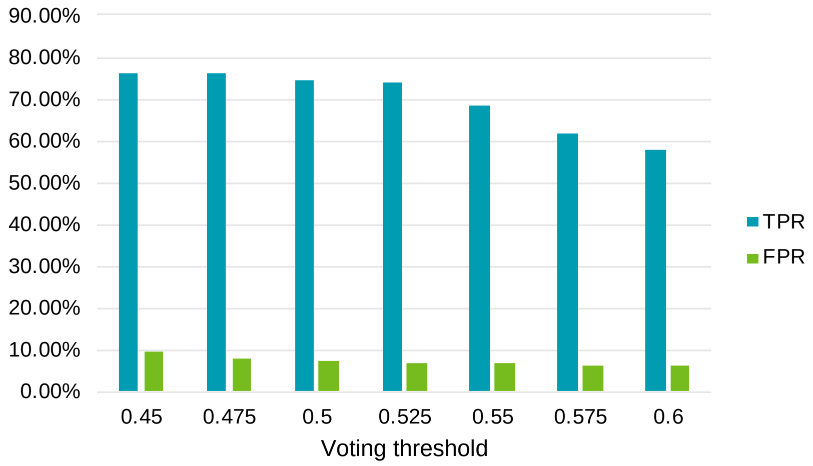

SUPPLEMENTARY FIGURE 5B. EXTRA TREE SLIDE-LEVEL PERFORMANCE BY VOTING THRESHOLD

SUPPLEMENTARY FIGURE 6: SHAPLEY VALUE OF EXTERNAL VALIDATION COHORT

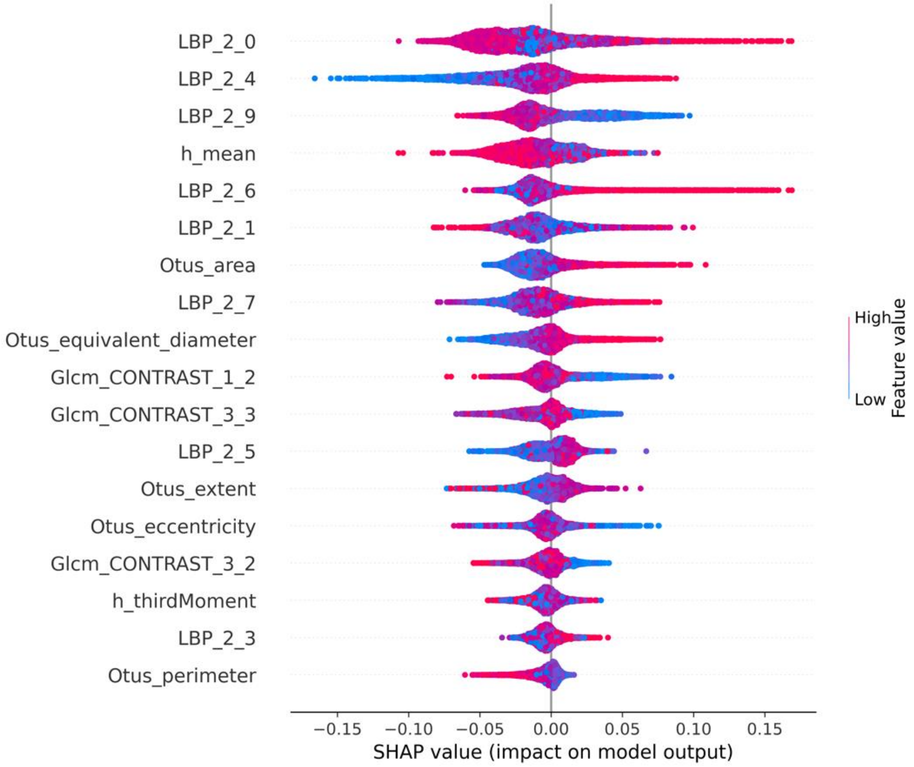

SHAPLEY value shows the relationship between the 18 histomic features and the outcome of CSFR with mesalamine alone at one year on external validation cohort. Positive SHAP-values (x-axis) are indicative of clinical remission, and negative value of non-remission
